# Nutritional Impacts of Minimum Unit Pricing for Alcohol: Are there unintended diet consequences?

**DOI:** 10.1101/2022.12.12.22283347

**Authors:** Attakrit Leckcivilize, Stephen Whybrow, Ni Gao, Lynda McKenzie, Daniel Kopasker, Paul McNamee, Anne Ludbrook

**Affiliations:** Health Economics Research Unit, University of Aberdeen, UK; Rowett Institute, University of Aberdeen, UK; Health Systems Collaborative, Nuffield Department of Clinical Medicine, University of Oxford, UK; Social & Public Health Science Unit, University of Glasgow, UK

## Abstract

**Background:** In 2018, Scotland introduced a Minimum Unit Pricing (MUP) policy to remove very-low-cost alcoholic drinks from the market and reduce adverse social and health-related impacts of excessive alcohol consumption. Any increased spending on alcohol may reduce spending on food, and adversely impact diet quality.

**Objectives:** To estimate the effect of MUP on dietary energy, nutrients and diet quality.

**Methods:** Analyses were conducted on household level purchase data, collected by Kantar Worldpanel (KWP) over 53 weeks before and 54 weeks after the introduction of MUP, from 1987 households in Scotland and 6064 households in the north of England. Energy and food purchases were adjusted to per adult-equivalent values after accounting for estimated unavoidable food waste. Difference-in-differences analyses were conducted for energy, energy density, Diet Quality Index, and foods and nutrients that are relevant to the Scottish dietary goals. The Poisson pseudomaximum likelihood regression model with household fixed-effects was used, with estimates adjusted for age of main shopper, household composition, duration of KWP participation, total spending on non-food items, and month of the year. The effects of area level deprivation and levels of alcohol purchase were also explored.

**Results:** The introduction of MUP in Scotland led to a 1.6% (95% Confidence Interval (CI) 0.02% - 3.16%) reduction in sugar purchase or 8 grams per adult-equivalent per week. This reduction was partly a result of a 16.6% (95% CI 7.15% - 25.96%) reduction in sugar from alcoholic drinks purchased. No significant associations were found between MUP and energy, energy density, other nutrients or diet quality. Households from more deprived areas, or with greater alcohol purchases, had greater levels of sugar reduction from alcohol.

**Conclusions:** The introduction of MUP in Scotland was associated with small, but beneficial, statistically significant reductions in the purchase of sugar. There was no significant change in overall diet quality.

## Introduction

Excessive consumption of alcohol in many countries is a major source of health problems including cancer and cardiovascular diseases (World Health Organisation 2019). Scotland has a more significant problem with alcohol misuse and higher alcohol related harms than other parts of the UK (Office for National Statistics 2020). For example, Scotland had higher alcohol-specific death rates (21.5 deaths per 100,000 persons) compared with England and Wales in 2020 (13.0 and 13.9 deaths per 100,000 persons) (Office for National Statistics 2021). In the past decades, a broad range of policies, including banning multi-buy price discounts and some promotional offers, has been introduced to tackle Scotland’s alcohol related problems. However, these policies could be made less effective by a wider use of simple price reductions.

On 1^st^ May 2018, Minimum Unit Pricing (MUP) for alcohol was introduced in Scotland. Scotland became the first country to set a strength-based floor price for alcohol (Beeston et al 2020). The legislation aimed to reduce alcohol consumption and adverse social and health impacts from alcohol-related harms by imposing a minimum price of 50 pence per unit (8g) of pure alcohol (NHS Health Scotland 2019). The policy will lapse after six years (2018 to 2024), unless it is extended by the Scottish Parliament following a report on the impact of MUP being led by Public Health Scotland. This report willconsider not only alcohol consumption but also other health and social costs and benefits, including the potential impact on household food expenditure and diet quality.

Evidence suggests increasing alcohol intake is associated with higher energy intake (e.g. Breslow et al. 2010, 2013; Grech et al. 2017; Kwok et al. 2019; Fong et al. 2021) and lower diet quality score (Breslow et al. 2013; Parekh et al. 2021). Moreover, frequent heavy drinking is associated with lower carbohydrate intakes (Cummings et al. 2020). Changes in the relative prices between some, previously cheap, alcoholic drinks and other food and drink purchases could lead to reductions in diet quality if increased spending on alcoholic drinks displaces healthier foods. Further, MUP mainly affects the price of alcohol sold in supermarkets, convenience stores and other premises licensed to sell alcohol for consumption off the premises (off sales)^1^. Off sales alcohol accounts for the majority of alcohol purchased in the UK (72.5% by volume in Scotland in 2017) (Richardson and Giles 2021). From an economic perspective, given a constrained budget, and with off sales alcohol frequently purchased in the same basket of goods as food, an increase in expenditure on alcohol may affect expenditure on food through the complementarity between these two products (Moore et al. 2021).

Previous research has shown that there was a small, statistically significant reduction in food expenditure caused by MUP and a non-significant reduction in total volume of food purchased (Kopasker et al 2022). This study, for the first time, examines the effect of MUP for alcohol on the nutritional components of food purchased for consumption at home by households. Using large-scale household consumer panel data collected by the UK Kantar Worldpanel (KWP), we focus on energy intake, dietary energy density, intakes for fruit and vegetables, fibre, oil-rich fish, total carbohydrate, red and processed meat, salt, percentage energy intake from fat, saturated-fat, trans-fat, and free-sugars, and diet quality index (DQI). Using the fact that MUP is implemented in Scotland rather than in England, we compare the nutritional components of food purchases and diet quality 12-months before and after the implementation of MUP between Scotland (treatment group) and north of England (control group). Our findings contribute to the wider evaluation of MUP and add to the literature on the nutritional relationship between food and alcohol.

## Method

### Data

We used large-scale household consumer panel data collected by the UK KWP. These data include weekly purchases on all food and drink brought into the home among panel members; items such as restaurant meals, takeaway food and on-premises alcoholic drinks are not included. Households used a handheld scanner device provided by KWP to scan a product’s bar-code. Non-bar-coded items (e.g. fruit and vegetables) were also recorded. The price of each purchased product was collected using households’ till receipts. For each recorded product purchased, the data include the description of the product, type of product (e.g. bread), quantity (weight or volume), and amount of money spent. For purchases of alcoholic drinks, the number of units of alcohol for each purchase was calculated from volume purchased and alcohol content (alcohol by volume (ABV), where one unit of alcohol = 8g pure alcohol). The ABV was detailed in the product description for most products, but where absent, a standard ABV value was used according to the type of alcoholic product (e.g. 40% for whisky).

#### KWP nutrient data

We focused on energy, fruit and vegetables, fish, total carbohydrate, red and processed meat, sugar, sugar excluding alcohol, sugar including alcohol only, fat, saturated fat, salt, fibre, energy density (kcal/100g) and Diet Quality Index (DQI). Nutritional information was collected by KWP from product labels, food composition tables, or product group averages. Composite foods were disaggregated to estimate the proportion of foods relevant to the Revised Dietary Goals for Scotland, such as fruit, vegetables and oil rich fish (Whybrow et al. 2018b). Detailed definitions of each nutrient and disaggregation were described in Whybrow et al. 2018b.

Energy density (kcal/100 g) of the food purchased was calculated from the contribution of all food and milks, but excluded all drinks (tea, coffee, water, fruit juices, squashes, sugar-containing drinks and artificially sweetened drinks). This is the same method as used in setting the Revised Dietary Goals for Scotland (Scottish Government 2016) and others (World Cancer Research Fund / American Institute for Cancer Research 2007; Wrieden et al. 2015).

A Diet Quality Index (DQI) was calculated for each household from each week’s food and drink purchases to allow comparison against the Revised Dietary Goals for Scotland (Scottish Government 2016). The “score” for diet quality ranges from 0%, to 100% if all the dietary goals are achieved. The DQI is broadly based on that developed by Barton et al. (2017) and modified for household purchase data (Whybrow et al. 2018a). KWP purchase data are recorded per household, whereas the dietary goals are set per person, and goals for some nutrients differ by age. To account for differing household composition, equalized household values were used to give per adult-equivalent values for food and drink purchases that were comparable to the dietary goals, by assuming that food and drink purchases are consumed by household members *pro rata* to estimated energy requirements. The total DQI score was calculated as the sum of the individual DQI components (see Supplemental Table 1).

#### Socioeconomic data

Socioeconomic information included household composition (age and gender of each household member, collected weekly), whilst socio-economic information for the main shopper (social-class, employment status), together with annual household income, were also available at less frequent intervals. For household location, KWP provides a Scottish Neighbourhood Statistics Data Zone identifier (The Scottish Executive 2004) for Scotland and a postcode within the TV Broadcasting Audience Research Board areas for the north of England consisting of Border England, North East, North West, and Yorkshire (Sky Media 2016). The Scottish Data Zone identifier is then used to classify households in Scotland into quintiles by level of neighborhood deprivation using the Scottish Index of Multiple Deprivation (SIMD) 2020 (Scottish Government 2020) (where 1^st^ quintile is the most deprived).

#### Sample size

We used data from the week ending the 30th of April 2017 to the week ending the 12th of May 2019, equivalent to 53 weeks before the MUP implementation and 54 weeks after that. As KWP members report their purchases for periods ranging from a few months to many years, only households with at least one observation week in both the pre-MUP and post-MUP periods are included in the analysis. The KWP data used for these analyses includes 1987 participating households in Scotland and 6064 households in the north of England.

### Statistical analysis

We employ a difference-in-differences (DID) framework to compare nutritional components of foods and quality of diet purchased by households in Scotland (treatment group) with those purchases made by households in the adjacent north of England (control group) before and after the implementation of MUP in Scotland. This DID design sets up a treatment-control comparison where the changes in north of England purchases are assumed to be a counterfactual for the changes in Scotland had the MUP policy not been introduced in Scotland.

Many of the outcome variables have a positively skewed distribution with many zeros concentrated in the left tail of the distribution, therefore, the Poisson pseudomaximum likelihood (PPML) regression model with household fixed-effects was used to perform the DID analysis since it makes minimal assumptions about the distribution of the data. Estimation was performed with Stata 14.2 using the user-written command, *ppmlhdfe* for estimating Poisson pseudomaximum likelihood with high-dimensional fixed effects (Correia et al 2020), which allows multiple sources of heterogeneity to be controlled for. The treatment effect is estimated from the coefficient of an exposure dummy variable (post-MUP in Scotland). To compute the effect size as a percentage change, the coefficient was transformed as exp^coefficient-1^.

As the sample of households from the north of England was substantially larger than the Scottish sample, with potential heterogeneities in observable characteristics between the two samples prior to the implementation of MUP, we applied entropy balancing to re-weight and balance inequalities in the means and variances of the observable characteristics. This method is a data pre-processing method to achieve covariate balance in observational studies by reweighting the control group (Hainmueller 2012). These balance improvements can reduce model dependence for the subsequent estimation of treatment effects (Hainmueller and Xu 2013). The weight was generated so that it minimizes the entropy distance metric of selected covariates subject to a set of balance constraints equating the moments of the covariate distribution (mean and variance) between the treatment and the reweighted control group. The Stata user-written package *ebalance* (Hainmueller and Xu 2013) was used to compute unit weights for re-weighting selected covariates before MUP in the control groups.

Using PPML, we first examined the average effect of MUP on food purchase and diet quality. As MUP may not have equal effects on different populations, we then examined heterogeneous effects of the MUP on households living in neighborhoods with different levels of deprivation, measured by SIMD, and on those with different levels of purchase of alcohol (up to 14 units versus over 14 units per adult per week). An interaction term is added between the treatment effects dummy and living in the top two quintiles of the SIMD (the least deprived areas) and between treatment effects dummy and purchasing higher level of alcohol purchase (over 14 units per adult per week).

## Results

### Sample balance at baseline

Table 1 compares the baseline characteristics (before MUP) between Scotland and north of England (unweighted and weighted). After entropy weighting, there were no significant differences in baseline characteristics between Scotland and weighted north of England. The average age of household shoppers was 52 years old, with over 70% women. Around 40% household shoppers worked over 30 hours per week, with income mainly concentrated between £10,000 to £29,999.

**Table 1.** Baseline characteristics of the KWP panel members before the introduction of MUP for Scotland and north of England.

|  | Scotland | North of England<br>(unweighted) | North of England<br>(weighted) |
| --- | --- | --- | --- |
| **Age** | 52.47 | 51.91 | 52.50 |
| **Household size** | 2.48 | 2.68 | 2.49 |
| **Having children (yes)** | 0.29 | 0.32 | 0.28 |
| **Main shopper (female)** | 0.71 | 0.73 | 0.70 |
| **Employment** |  |  |  |
| under 8 hours | 0.02 | 0.02 | 0.02 |
| 8-29 hours | 0.17 | 0.19 | 0.17 |
| full-time education | 0.00 | 0.00 | 0.01 |
| over 30 hours | 0.42 | 0.39 | 0.42 |
| retired | 0.26 | 0.27 | 0.26 |
| not working | 0.11 | 0.11 | 0.11 |
| unemployed | 0.02 | 0.02 | 0.02 |
| **Annual household income** |  |  |  |
| £0-£9999 | 0.07 | 0.07 | 0.07 |
| £10000-£19999 | 0.23 | 0.23 | 0.22 |
| £20000-£29999 | 0.22 | 0.23 | 0.22 |
| £30000-£39999 | 0.18 | 0.18 | 0.18 |
| £40000-£49999 | 0.12 | 0.12 | 0.12 |
| £50000-£59999 | 0.08 | 0.08 | 0.08 |
| £60000-£69999 | 0.05 | 0.04 | 0.05 |
| £70000+ | 0.06 | 0.06 | 0.06 |
| **Social class** |  |  |  |
| AB | 0.21 | 0.21 | 0.21 |
| C1 | 0.38 | 0.39 | 0.39 |
| C2 | 0.16 | 0.18 | 0.17 |
| D | 0.15 | 0.13 | 0.15 |
| E | 0.10 | 0.09 | 0.09 |
| **Household location: rural** | 0.21 | 0.14 | 0.21 |
| **Sample weight** | 1.00 |  | 0.32 |
| **Number of households** | 1987 | 6064 | 6064 |

\*All values were averaged before MUP (May-2018). There is no significant difference between sample of Scotland and weighted sample of England.

### Comparison of nutritional components and quality of diet

Table 2 summarizes the average nutritional components of food and DQI in Scotland and north of England, before and after MUP. These values suggest that the households in Scotland and north of England were similar in terms of intake of nutritional components, both before and after MUP. In terms of differences, considering the pre MUP values, the largest proportional difference related to sugar, where sugar purchase from all sources, and sugar purchase excluding alcohol, was 5g and 5.1g higher per adult equivalent per day in Scotland pre MUP.

**Table 2.** Mean (s.d.) daily energy, nutrients and foods available for consumption per adult equivalent per day (or per week for oil-rich-fish), in Scotland and north of England.

| Outcome variables | Before MUP |  |  | After MUP |  |  |
| --- | --- | --- | --- | --- | --- | --- |
|  | Scotland | North of England | Difference | Scotland | North of England | Difference |
| Energy (kcal) | 2962<br>(2090) | 2947<br>(2101) | 15 | 2966<br>(2130) | 2992<br>(2155) | -26 |
| Energy density (kcal/100g) | 197.0<br>(140.6) | 197.2<br>(142.5) | -0.20 | 198.4<br>(144.2) | 200.5<br>(146.2) | -2.10 |
| Fruit and vegetables (g) | 405<br>(387) | 400<br>(367) | 4.6 | 402<br>(386) | 404<br>(371) | -1.4 |
| Oil rich fish (g per week) | 36.6<br>(136.5) | 41.6<br>(144.5) | -4.9 | 36.7<br>(131.8) | 43.8<br>(147.2) | -7.1 |
| Red and processed meat (g) | 175<br>(230) | 175<br>(236) | 0.3 | 176<br>(239) | 178<br>(241) | -2.1 |
| CHO (g) | 299<br>(215) | 298<br>(215) | 1.8 | 297<br>(216) | 299<br>(216) | -1.9 |
| Sugar (g) | 79<br>(84) | 74<br>(79) | 5.0 | 76<br>(80) | 73<br>(78) | 2.8 |
| Sugar excluding alcohol (g) | 78<br>(83) | 73<br>(78) | 5.1 | 75<br>(80) | 71<br>(77) | 3.2 |
| Sugar including alcohol only (g) | 1.2<br>(5.37) | 1.3<br>(4.72) | -0.1 | 1<br>(3.75) | 1.3<br>(4.75) | -0.3 |
| Fat (g) | 132<br>(110) | 130<br>(109) | 2.0 | 134<br>(113) | 133<br>(113) | 1.1 |
| Saturated fat (g) | 52<br>(44) | 51<br>(44) | 1.1 | 52<br>(46) | 51<br>(45) | 0.9 |
| Salt (g) | 10.0<br>(9.06) | 9.9<br>(9.16) | 0.0 | 9.9<br>(9.14) | 9.9<br>(9.12) | 0.0 |
| Fibre (g) | 22<br>(16) | 22<br>(17) | -0.6 | 22<br>(17) | 23<br>(17) | -0.7 |
| Diet Quality Index (%) | 37.1<br>(16.2) | 38.1<br>(16.3) | -1.03 | 37.2<br>(16.2) | 38.3<br>(16.4) | -1.10 |

Supplemental Figure 1 also illustrates the trends of nutritional components of food and DQI during the pre-MUP and post-MUP periods for Scotland and north of England. Overall, trends of all outcomes are similar between Scotland and north of England.

### Average effect of MUP on food purchase and DQI

Figure 2 illustrates the effect of MUP on percentage change in food purchase and DQI. Compared with the north of England, MUP has no significant effect on most nutrient intakes and DQI. However, MUP has a significant effect on sugar purchase. Given that sugar is found in some alcoholic drinks, which could be affected directly by MUP, sugar intakes are classified into sugar from all sources, sugar from all sources excluding alcoholic drinks, and sugar from alcoholic drinks only. MUP significantly reduces total sugar purchase by 1.6% per week in Scotland, or approximately 8 grams per individual per week. MUP significantly reduces purchase on sugar from alcoholic drinks by 16.6% per week, or approximately 1.4 grams less per individual per week. MUP significantly reduces purchase on sugar excluding alcoholic drinks by 1.4% per week, or approximately 7 grams less per individual per week. Table 3 extends Figure 2 by showing the exact coefficient estimates, standard errors, calculated percentage changes, and level changes as calculated from the mean values and percentage change estimates.

**Figure 2.**
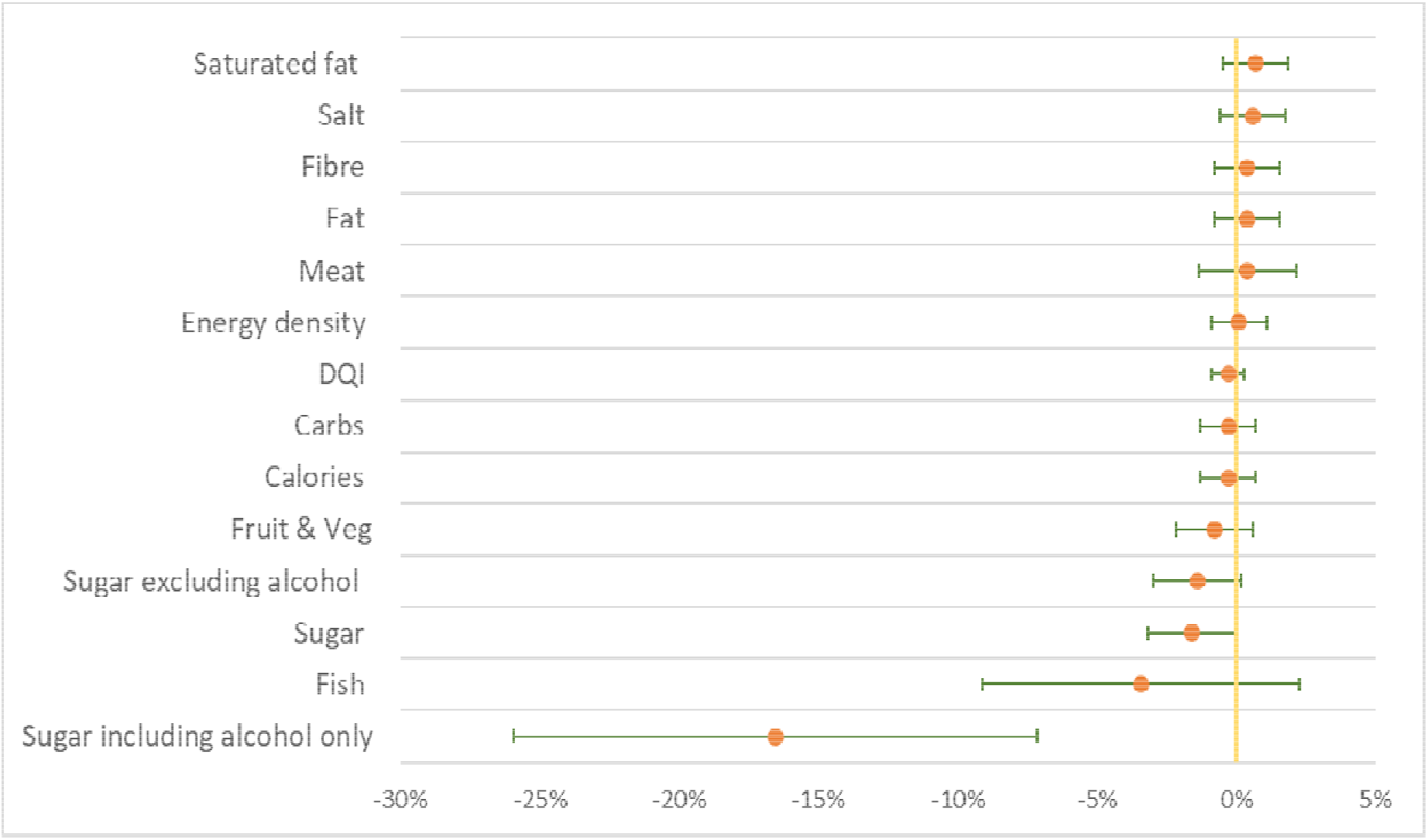
The effect of MUP on food purchase and DQI (percentage change with 95% confidence interval)

**Table 3.** Estimated treatment effects of the MUP in Scotland on energy, food and nutrient intakes and DQI score. Values are mean daily amounts per adult equivalent.

|  | Energy<br>(kcal) | Energy<br>density<br>(kcal/100<br>g) | Fruit & Vegeta-<br>bles (g) | Oil-rich-fish<br>(g/week) | Red and<br>processed<br>meat (g) |
| --- | --- | --- | --- | --- | --- |
| Mean | 2968 | 199 | 402 | 41.2 | 176 |
| Coefficients from PPML | -0.003 | 0.001 | -0.008 | -0.035 | 0.004 |
| Standard error | (0.005) | (0.005) | (0.007) | (0.029) | (0.009) |
| Percentage change (%) | -0.3 | 0.1 | -0.8 | -3.4 | 0.4 |
| Level change | -8.9 | 11.8 | -3.2 | -1.4 | 0.7 |

|  | CHO (g) | Sugar (g) | Sugar excluding<br>alcohol (g) | Sugar including<br>alcohol only (g) | Fat (g) |
| --- | --- | --- | --- | --- | --- |
| Mean | 298 | 74.3 | 73.1 | 1.25 | 131.8 |
| Coefficients from PPML | -0.003 | -0.016** | -0.014* | -0.181*** | 0.004 |
| Standard error | (0.005) | (0.008) | (0.008) | (0.048) | (0.006) |
| Percentage change (%) | -0.3 | -1.6 | -1.4 | -16.6 | 0.4 |
| Level change | -0.9 | -1.2 | -1.0 | -0.2 | 0.5 |

|  | Saturated<br>fat (g) | Salt (g) | Fibre (g) | DQI (%) |
| --- | --- | --- | --- | --- |
| Mean | 51.3 | 9.91 | 22.4 | 37.9 |
| Coefficients from PPML | 0.007 | 0.006 | 0.004 | -0.003 |
| Standard error | (0.006) | (0.006) | (0.006) | (0.003) |
| Percentage change (%) | 0.7 | 0.6 | 0.4 | -0.3 |
| Level change | 0.4 | 0.1 | 0.1 | -0.1 |

Percentage change was calculated using coefficients from PPML: $\exp(\text{coefficient})-1$ ; level change was calculated based on the mean of outcomes and percentage change: $\text{mean} \times [\exp(\text{coefficient})-1]$ . For example, level change for energy is calculated as $2968 \times [\exp(-0.003)-1] = -8.9$ kcal per person per day.
All analysis is adjusted for social economic factors including age, total people in household, whether having children, spending on non-food categories, and month dummies.
\*\*\*p<0.01; \*\*p<0.05; \*p<0.1.

### Heterogeneous effect of MUP on food purchases and DQI

MUP may have a heterogenous effect on households with different socioeconomic status. Supplemental Table 2 compares the effect of MUP between households from the bottom three quintiles of SIMD and top two quintiles of SIMD. Overall, MUP has no significant heterogenous effect among the two groups across most of the measures. However, households from the bottom three quintiles of SIMD reduce purchase on sugar from alcohol significantly more than those from the top two quintiles of SIMD. Also, households from the bottom three quintiles of SIMD reduce purchase of fish significantly more than those from top two quintiles of SIMD.

Supplemental Table 3 compares the effect of MUP between households with high level of alcohol purchase (>14 units per adult per week) and moderate level of alcohol purchase (1-14 units per adult per week). Again, amongst most measures, MUP has no significant heterogenous effect among the two groups. However, households with high level of alcohol purchase reduce purchase of sugar from alcohol significantly more than those with moderate level of alcohol purchase.

## Discussion

Our findings demonstrate that the introduction of MUP for alcohol was not associated with significant adverse effects on nutrition in Scotland. There were no significant associations with changes in overall diet quality or nutrients except for sugar. The significant effect of MUP policy on reduction in sugar intakes is potentially beneficial and is found for sugar from all sources and sugar from alcohol only. Moreover, in the subgroup analysis, a reduction in sugar is observed for households in the more deprived areas.

The effect size of the policy on sugar purchased may appear modest at approximately 8 grams per adult equivalent per week on average, especially given the findings that there is no improvement in the diet quality score for sugar intakes. Yet this estimated reduction is non-trivial compared to the impacts of policies specifically targeting sugar consumption in the UK, such as the Soft Drinks Levy introduced prior to MUP. An evaluation of the levy, also using Kantar data, estimated a reduction in sugar purchased of 29.5g per household per week (Pell et al 2021). Previous voluntary action by Public Health England and the food industry to reduce sugar content had not reduced sugar purchased per person (Public Health England 2020).

The impact of MUP on the purchase of added sugar from alcoholic drinks is negative and significant, both across the entire population and within particular sub-groups, notably those purchasing higher levels of alcohol and those living in more deprived areas. These findings not only confirm a significant drop in alcohol purchases in Scotland post-MUP reported in previous studies (O’Donnell et al 2019; Anderson et al 2021; Public Health Scotland 2021) but also support the heterogeneous impacts found in those studies (O’Donnell et al 2019; Anderson et al 2021), where the high purchase households decreased their alcohol purchasing significantly more. Furthermore, a report by Public Health Scotland 2021 showed that the biggest reduction in consumption following MUP was of cider and perry, which are drinks with a high sugar content.

The health impacts of reduced sugar consumption appear considerable (Amies-Cull et al. 2019). Using data from the National Diet and Nutrition Survey, the authors estimate that a 5% reduction in sugar intake from baseline levels would be projected to produce NHS cost savings of £124 million for males and £162 million for females, and increased health benefits through additional Quality Adjusted Life Years gained of between 23,874 (males) and 27,855 (females), measured over a 10 year period. The effects were generated through a reduction of 19kcal/day for adults and a weight reduction of between 1.5-1.8kg. The health benefits accrued were generated from avoided cases of cardiovascular disease, stroke, diabetes, cirrhosis and cancer, with the largest cases avoided being generated from diabetes. However, the modelling does not take account of consumer response in the form of additional consumption of other products, or indeed additional consumption of those products that were re-formulated. In addition, the estimates rely on self-reported cross-sectional associations between risk factors and disease. Therefore, it is possible that the estimates are upper limits of the potential impacts.

Regarding fish purchases, our finding of reductions in purchase amongst the subgroup of people from more deprived households is consistent with a previous study that used the same KWP data and found a significant reduction across the population in the volume of fish purchases following MUP(Kopasker et al 2022). It is difficult to pinpoint reasons why MUP would be associated with such a change, which suggests this finding is either artefactual or driven by unobservable factors. In addition, the data for fish are highly skewed and the consumption of oily fish by the most deprived groups was reported to be half the level of less deprived groups prior to MUP being introduced (Food Standards Scotland 2018).

A strength of this study is the use of a robust DID design and a large dataset with detailed dietary information. The parallel trends assumption holds in our sample prior to the introduction of MUP (shown in Supplemental Figure 1). Although the Soft Drinks Industry Levy (sugar tax) was implemented in the whole UK less than a month before MUP came into effect, the sugar tax affected both Scotland and the north of England equally.

There are some limitations of our study due to the nature of the KWP data. First, we have no information on purchases consumed outside the home; however, we are not aware of any changes other than MUP that would have a differential impact on this in Scotland compared with the north of England. Secondly, we implicitly assume that households consume all food and drinks they bought in that particular week. This is unlikely because some food and drinks might be bought and kept longer for future consumption and some could end up as food waste, particularly among fresh products. Indeed, systematic reviews (e.g. Schanes et al 2018) indicate that people often follow routines of buying more food than needed and over-provisioning of food is one of the main reasons for food waste in private households.

Although we are aware that wasted food causes sizable nutrient loss (Spiker et al 2017), purchasing data such as KWP do not provide any information on food waste in the households. However, with weekly panel data collection, the issues of time inconsistency between purchase and consumption should be mitigated because households in Scotland and the north of England should have similar and stable patterns in food and drink consumption (in terms of days between purchasing and consuming) after their purchases.

It has also to be recognised that the sample provided by the KWP, whilst representative of the population, does not fully cover the heaviest purchasers of alcohol. In a qualitative study of people who are drinking at harmful levels (more than 35 units for women (50 units for men) per week) or dependent on alcohol, some have reported reducing expenditure on food and, in some cases, using food banks to access free food (Holmes et al 2022). Equally, some of those in the study reported being more likely to seek treatment for their drinking.

One remaining concern is the effect of MUP on the shares of off-trade sales and on-trade sales (e.g. pubs and clubs) of alcohol in Scotland. Although the MUP applied to all types of alcohol sales in Scotland, it could narrow the price difference between on-trade sales and off-trade sales because the price of on-trade sales tended to be higher before the MUP introduction in May 2018. A reducing gap in Scotland could encourage a substitution from off-trade purchases towards on-trade consumption, and hence could partially reverse the negative impact of MUP on sugar intakes, especially sugar from alcoholic drinks. However, aggregated level data published by Public Health Scotland (2021) depicts similar trends in on-trade sales of alcohol (litres of pure alcohol per adult) between Scotland and England during 2018 and 2019, decreasing slightly from the 2017 level. Thus, our estimated effects should not be seriously altered and can be considered a conservative upper bound of the reduction of sugar intakes arising from the policy.

In conclusion, the analysis presented here suggests that the introduction of MUP had little significant effect on nutrition from food purchased to eat at home, except for a beneficial effect on sugar consumption. The potential for further impact should, however, continue to be considered as part of any future review of changes to MUP policy.

## Data Availability

Kantar Worldpanel data are not publicly available but can be purchased from Kantar Worldpanel (http://www.kantarworldpanel.com). The authors are not legally permitted to share the data used for this study.

## Acknowledgements

The study was funded by a research grant from the Chief Scientist Office (CSO) of the Scottish Government Health and Social Care Directorates, HIPS Grant Number HIPS/19/01. The Health Economics Research Unit is also partly funded by the CSO. The views expressed in the paper are those of the authors only and not those of the funder.

## Online Supplementary

**Supplemental Table 1:** Components of the Diet Quality Index and scoring criteria.

| Component | Scottish dietary goal | Scoring criteria |
| --- | --- | --- |
| Energy density | Average energy density of the diet to be lowered to 125 kcal/100 g | $\leq 125 \text{ kcal/100 g} = 10$<br>$> 125 \text{ kcal/100 g} = 0$ |
| Fruits and vegetables | Average intake of a variety of fruits and vegetables to reach > 400 g per person per day | Scaled from 0 for 0 g per day to a maximum of 10 for $\geq 400 \text{ g per day}$ |
| Oil-rich fish | Oil-rich fish consumption to increase to one portion per person (140 g) per week (= 20 g per day) | Scaled from 0 for 0 g per day to a maximum of 10 for $\geq 20 \text{ g per day}$ |
| Red and processed meat | Average intake of red and processed meat to be pegged at around 70 g per person per day | $\leq 70 \text{ g per day} = 10$<br>$> 70 \text{ g per day} = 0$ |
| %Energy carbohydrate | Total carbohydrate to be maintained at an average population intake of approximately 50% of total dietary energy with no more than 5% total energy from free sugars | $\geq 40\% \text{ and } \leq 60\% = 10$<br>$< 40\% = 0$<br>$> 60\% = 0$ |
| %Energy sugar | Average intake of free sugars, not to exceed 5% of total energy in adults and children over two years | $\leq 5\% \text{ energy} = 10$<br>$> 5\% \text{ energy} = 0$ |
| %Energy total fat | Average intake of total fat to reduce to no more than 35% food energy | $\leq 35\% \text{ energy} = 10$<br>$> 35\% \text{ energy} = 0$ |
| %Energy saturated fat | Average intake in saturated fat to reduce to no more than 11% food energy | $\leq 11\% \text{ energy} = 10$<br>$> 11\% \text{ energy} = 0$ |
| %Energy trans fat | Average intake of trans fatty acids to remain below 1% food energy | Not scored<sup>a</sup> |
| Salt | Average intake of salt to reduce to 6 g per day | $\leq 6 \text{ g per day} = 10$<br>$> 6 \text{ g per day} = 0$ |
| Fibre | An increase in average consumption of AOAC fibre for adults (16+) to 30 g/day. Dietary fibre intakes for children to increase in line with SACN recommendations | Scaled from 0 for 0% of requirements to a maximum of 10 for $\geq 100\%$ of requirements |

<sup>a</sup>Scoring not possible because nutrient information is not collected by Kantar WorldPanel. AOAC: Association of Official Analytical Chemists; SACN: Scientific Advisory Committee on Nutrition.
**Note:** The total estimated energy requirement for each household was calculated by summing the requirements for adults and children, accounting for age and sex (Scientific Advisory Committee on Nutrition 2009). The estimated energy requirement of children under 2 years old was not included. A reference energy requirement for all adults was estimated as the average for 19–59 years old males and females (2223 kcal per day) (Scientific Advisory Committee on Nutrition 2009). This allowed food and nutrient intake to be estimated per adult equivalent from combined household purchases. For example, if a household of two elderly adults, with an estimated energy requirement of 4254kcal per day, purchased 300g of fruit in a week this would be $300 / 7 / (4254 / 2223) = 22.4\text{g}$ per adult equivalent per week.

**Supplemental Table 2:**
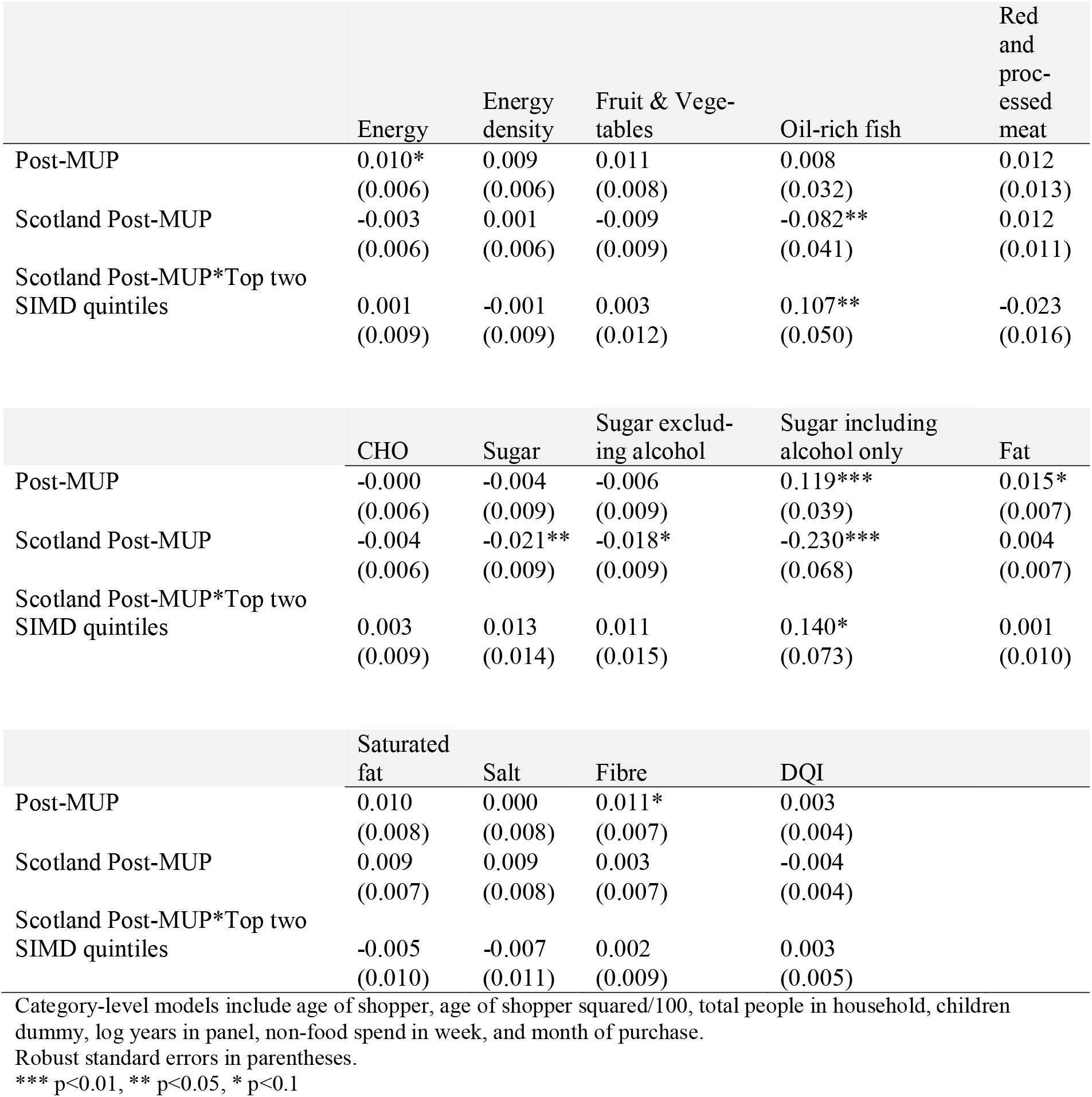
Heterogeneous treatment effects of the MUP in Scotland by deprivation level in the neighborhood area of households on nutrients intakes and DQI score (percentage change)

**Supplemental Table 3:** Heterogeneous treatment effects of the MUP in Scotland by level of alcohol purchase in the households on nutrients intakes and DQI score (percentage change)

|  | Energy | Energy density | Fruit & Vegetables | Oil-rich-fish | Red and processed meat |
| --- | --- | --- | --- | --- | --- |
| Post-MUP | 0.010*<br>(0.006) | 0.009<br>(0.006) | 0.011<br>(0.008) | 0.008<br>(0.032) | 0.012<br>(0.013) |
| Scotland Post-MUP | -0.002<br>(0.005) | 0.001<br>(0.005) | -0.008<br>(0.008) | -0.041<br>(0.031) | 0.006<br>(0.010) |
| Scotland Post-MUP*High alcohol purchase | -0.009<br>(0.014) | -0.002<br>(0.014) | 0.003<br>(0.019) | 0.038<br>(0.071) | -0.015<br>(0.021) |

|  | CHO | Sugar | Sugar excluding alcohol | Sugar including alcohol only | Fat |
| --- | --- | --- | --- | --- | --- |
| Post-MUP | -0.000<br>(0.006) | -0.004<br>(0.009) | -0.006<br>(0.009) | 0.118***<br>(0.039) | 0.015*<br>(0.007) |
| Scotland Post-MUP | -0.003<br>(0.006) | -0.018**<br>(0.008) | -0.017**<br>(0.008) | -0.050<br>(0.034) | 0.004<br>(0.006) |
| Scotland Post-MUP*High alcohol purchase | 0.002<br>(0.015) | 0.013<br>(0.023) | 0.027<br>(0.024) | -0.251***<br>(0.085) | -0.001<br>(0.016) |

|  | Saturated fat | Salt | Fibre | DQI |
| --- | --- | --- | --- | --- |
| Post-MUP | 0.010<br>(0.008) | 0.000<br>(0.008) | 0.011*<br>(0.007) | 0.003<br>-0.004 |
| Scotland Post-MUP | 0.007<br>(0.006) | 0.008<br>(0.006) | 0.004<br>(0.006) | -0.004<br>-0.003 |
| Scotland Post-MUP*High alcohol purchase | -0.005<br>(0.016) | -0.011<br>(0.020) | -0.004<br>(0.015) | 0.004<br>-(0.009) |

Category-level models include age of shopper, age of shopper squared/100, total people in household, children dummy, log years in panel, non-food spend in week, and month of purchase.
Robust standard errors in parentheses.
\*\*\* p<0.01, \*\* p<0.05, \* p<0.1

**Supplemental Figure 1.**
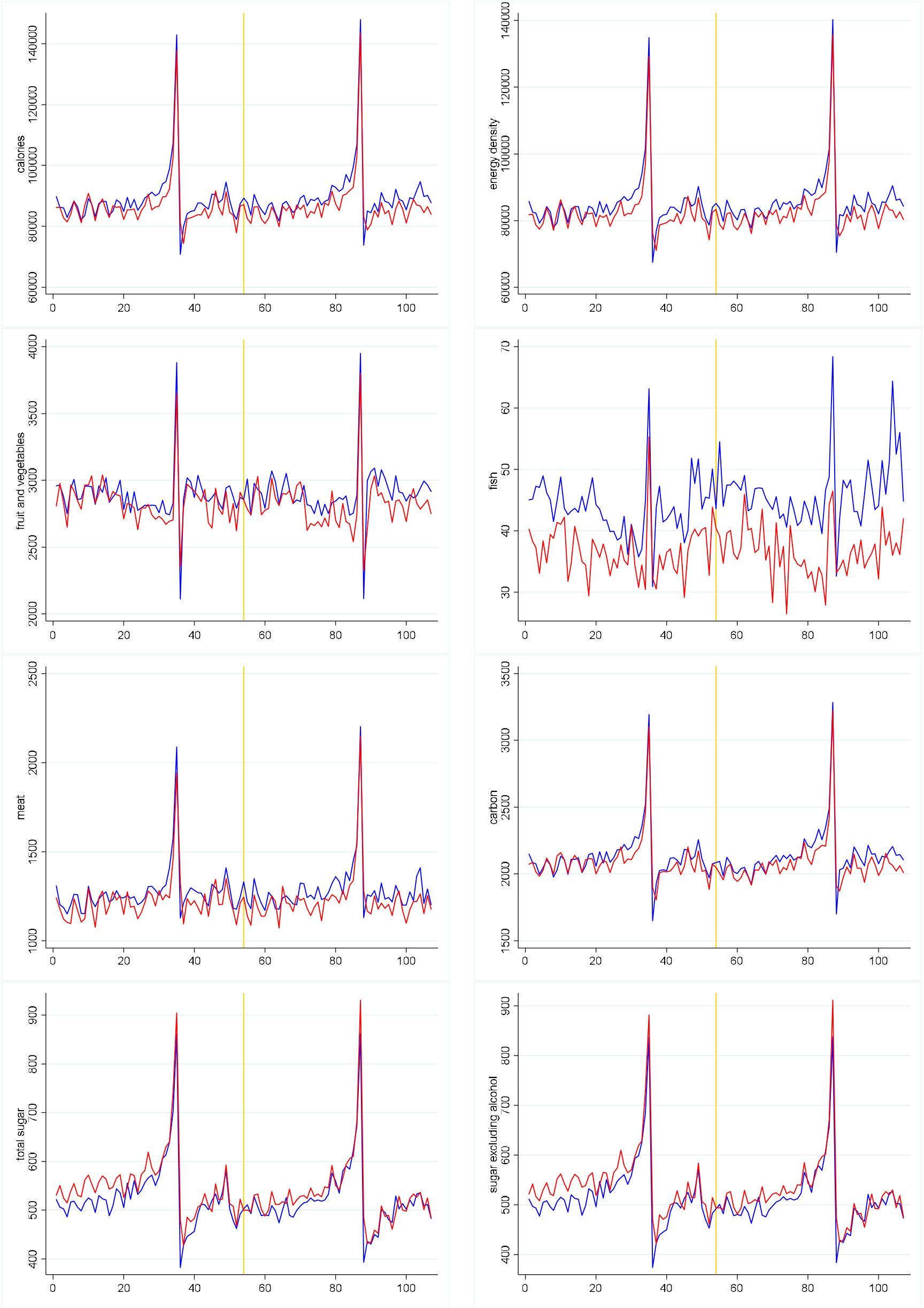
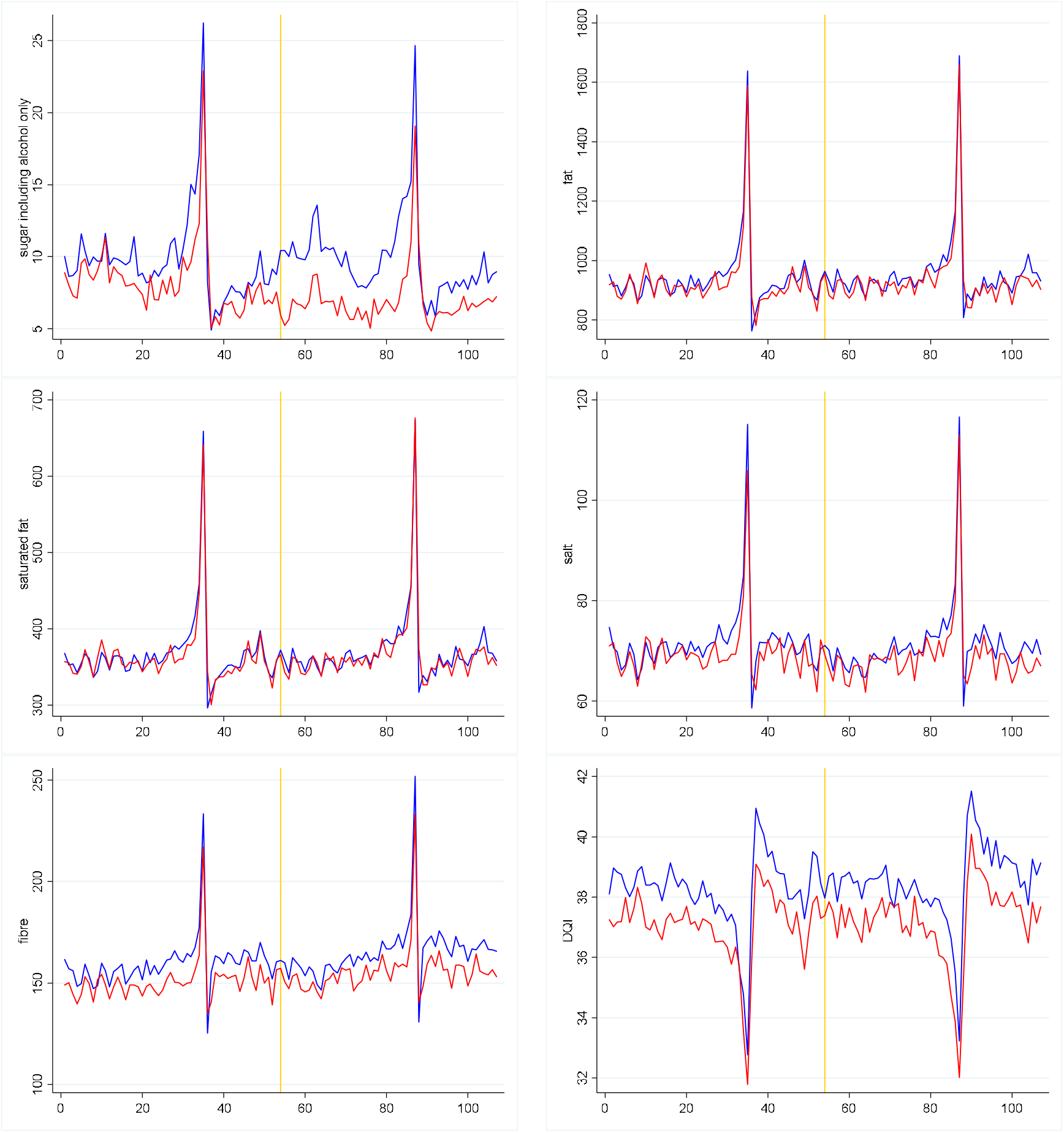
Trends of nutritional components of food and DQI between Scotland (red line) and North of England (blue line) *Yellow line is the time MUP implemented; X-axis is weeks before and after MUP; for Y-axis, see Table 2.

## Footnotes

1 Alcohol sold for consumption in bars and restaurants (on sales) would mostly be above the MUP threshold (i.e. 50 pence per unit), and therefore be unaffected by the legislation.

